# The effect of angiotensin converting enzyme inhibitors and angiotensin receptor blockers on death and severity of disease in patients with coronavirus disease 2019 (COVID-19): A meta-analysis

**DOI:** 10.1101/2020.04.23.20076661

**Authors:** S Ghosal, Jagat J Mukherjee, B Sinha, K Gangopadhyay

**Author notes:** Corresponding author: Samit Ghosal.

## Abstract

**Aims and Methods:** Effect of angiotensin converting enzyme inhibitors (ACEi) and angiotensin receptor blockers (ARB) on outcomes in patients with coronavirus disease 2019 (COVID-19) is uncertain. Available evidence is limited to a few retrospective observational studies with small number of patients. We did a meta-analysis to assess the effect of ACEi/ARB in patients with COVID-19 on severity of disease, risk for hospitalisation, and death compared to those not on ACEi/ARB. We searched the Cochrane library, PubMed, Embase, ClinicalTrial.gov and medRxiv for studies published until 25.04.2020. Inclusion criteria included all studies with patients with confirmed COVID-19 either taking, or not taking, ACEi/ARB. Depending on degree of heterogeneity, fixed or random effect model was selected to calculate effect size (Odds ratio).

**Results:** Six studies were eligible for this meta-analysis. These included 423 patients on ACEi/ARB, and 1419 not on ACEi/ARB. Compared to patients with COVID-19 not on ACEi/ARB, there was a statistically significant 43% reduction (OR 0.57, CI: 0.37–0.88, I^2^: 0.000) in the odds of death in those on ACEi/ARB. There was a statistically non-significant 38% reduction (OR: 0.62, 95% CI: 0.31–1.23, I^2^=70.36) in the odds of developing severe disease and 19% reduction (OR 0.81; 95% CI: 0.42–1.55, I^2^: 0.000) in the odds of hospitalisation among those on ACEi/ARB.

**Discussion:** It is safe to use ACEi/ARB in patients with COVID-19 requiring these medications for associated comorbidities. Although limited by confounding factors typical of a meta-analysis of retrospective observational studies, our data suggests that use of these medications may reduce the odds of death.

**Conclusion:** Our meta-analysis of the updated studies on SARS-CoV-2 reassures the medical fraternity on the use of and continuation of ACEi/ARB, supporting all recent recommendations.

**Evidence before this study:**

- The postulated dual role of angiotensin-converting enzyme (ACE) inhibitors (ACEi) and angiotensin receptor blockers (ARB) in patients with coronavirus disease 2019 (COVID-19) has created a dilemma for clinicians.
- On the one hand, there is speculation that by upregulating ACE2, ACEi/ARBs might increase the risk and severity of COVID-19.
- On the other hand, there is evidence that downregulation of ACE2 can mediate acute lung injury. Further evidence is urgently needed to guide clinicians in the use of ACEi/ARB in patients with COVID-19 with co-morbidities.

**What does this article add:**

- Our meta-analysis, which is the first to assess the effect of use of ACEi/ARB in patients with COVID-19, reports that use of ACEi/ARB statistically significantly reduced the risk of death, with a trend towards reduction in risk of severe disease and hospitalisation compared to those who were not on ACEi/ARB.
- Further information from on-going RCTs shall take time to fruition; in the interim, based on these findings, clinicians can safely continue to use ACEi/ARB in patients with COVID-19 with comorbidities.

**Review Criteria:**

- A web-based search was conducted using the Cochrane library, PubMed, Embase, ClinicalTrial.gov and medRxiv using specific keywords.
- Narrowing down of the citations was done based on full text availability and a set of pre-determined inclusion criteria.
- Meta-analysis was conducted on the pooled data comparing ACEi/ARB group versus the non-ACEi/ARB group on death, severity of disease and hospitalization using the CMA software version 3, Biostat Inc., Englewood, NJ, USA.
- Effect size was reported as odds ratio with a 95% confidence interval and the degree of heterogeneity of the pooled data.

**Message for the clinic:**

- There is no indication from present evidence to withhold or withdraw ACEi/ARB in patients with SARS-CoV-2.

## 1.0 Introduction

Coronavirus disease 2019 (COVID-19) is a rapidly unravelling pandemic infection caused by severe acute respiratory syndrome coronavirus 2 (SARS-CoV2). It is highly infectious, with a case fatality rate of 1% to 5% (1). Despite a wide variability in the prevalence and case fatality rates in the published case-series of COVID-19 from China and USA, it is now clear that patients with underlying pulmonary disease, cardiac disease, hypertension, diabetes mellitus (DM), and renal disease are not only more likely to contract SARS-CoV2 but also have higher morbidity and mortality rates (2, 3, 4). Angiotensin-converting enzyme inhibitors (ACEi) and angiotensin receptor blockers (ARB) are commonly used in patients with cardiac diseases, hypertension, many renal conditions, and DM, which directed attention towards the role of ACEi/ARBs in COVID-19, more so because SARS-CoV2 uses angiotensin-converting enzyme 2 (ACE2) as a co-receptor to enter cells (5).

ACE2, a homologue of angiotensin-converting enzyme 1 (ACE1), cleaves angiotensin II (Ang II), the main active peptide of the renin-angiotensin-aldosterone system (RAAS), to form angiotensin-(1–7) (6, 7). Angiotensin-(1–7) acts as a counter-regulatory peptide by attenuating the deleterious effects of angiotensin II, namely vasoconstriction, sodium retention and fibrosis. ACE2 is a transmembrane protein that is anchored to the apical surface of the cell. It can be cleaved and released into the circulation. ACE2 is expressed in the heart (cardiomyocytes, cardiac fibroblasts, coronary endothelial cells), kidney, testis, gastrointestinal system and type II alveolar cells in the lung (8).

Some, but not all, animal studies have shown that ACEi/ARBs upregulate ACE2 expression (9). This has led to the speculation that use of ACEi/ARB shall not only increase the risk but also the severity of SARS-CoV2 infection. However, unlike in animal models, in the few available human studies, the effects of RAAS inhibitors on ACE2 is far from clear (10, 11). Moreover, animal, and some human, studies have failed to establish the effects of ACEi or ARB on the expression of ACE2 in alveolar cells in the lung.

In contrast to the speculative detrimental effect of upregulation of ACE2 expression in COVID-19, elevated ACE2 levels has a number of beneficial effects. The conversion of Ang II to Ang-(1–7) ameliorates the harmful effects of Ang II, and instead mediates vasodilation and anti-inflammatory effects. Animal studies have suggested that downregulation of ACE2 may result in unopposed action of angiotensin II, that can mediate acute lung injury in viral infections (12), including SARS-CoV2 (13), which can be ameliorated by recombinant ACE2 (14) or RAAS blockade (15). Moreover, animal studies have demonstrated the protective effect of ACE2 on the myocardium (16).

This postulated dual role of ACEi/ARBs, based mostly on theoretical concerns and animal data, has created a dilemma for clinicians because many patients with co-morbidities like diabetes, hypertension, coronary artery disease and chronic kidney disease that increase susceptibility to, and a poor outcome in, SARS-Cov-2 infection, are on ACEi and ARBs. More evidence from the laboratory, and the clinics, are urgently needed to guide frontline physicians during this rapidly progressive pandemic with potential to cause massive devastation. Among these, retrospective analyses of outcomes in patients with COVID-19 who were on ACEi/ARBs can serve as a good evidence base. However, such analyses are limited at present due to very few retrospective case studies with outcomes in patients with SARS-CoV2 infection on ACEi/ARBs (17, 18, 19, 20, 21, 22); moreover, the number of patients included, and the number of outcomes of interest, are small in these individual studies to draw any meaningful conclusions. We therefore did a meta-analysis of the studies reported till 21.04.2020 on the outcomes in patients with COVID-19 on ACEi/ARBs to strengthen the evidence base by determining the benefit-risk ratio of these medications in patients with COVID-19.

## 2.0 Materials and methods

An electronic database search was conducted using the Cochrane library, PubMed, Embase, CT.gov and mdRxiv. Search keywords included “angiotensin receptor blockers”, “angiotensin converting enzyme inhibitors”, “ACE inhibitors”, “ARB”, “SARS”, “severe acute respiratory syndrome”, “COVID-19”, “SARS-CoV-2”, “mortality”, “death”, “complications”, “acute respiratory distress syndrome”, “mortality”, “death”. The search was conducted irrespective of any language barrier or time constraints. As a part of advanced screening references of identified citations were also screened for any additional information missed out in the screening process. [Figure 1]

**Figure 1.**
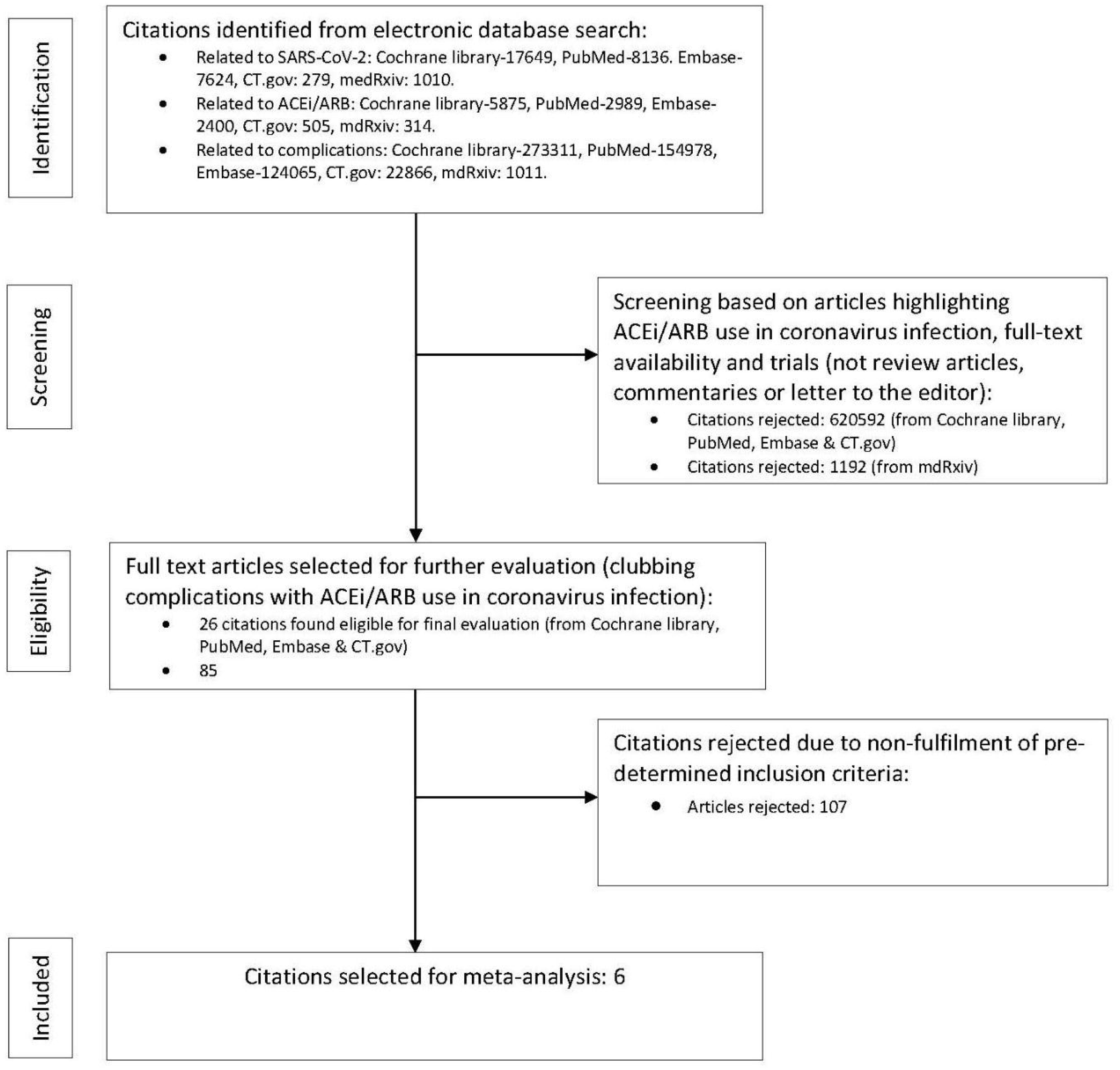

### 2.1 Inclusion criteria

A broad web-based search was conducted to identify all citations published, or in the pre-print stage. Having narrowed down the search tailored to the aims of our study, we finalised the citations to be included as part of the meta-analysis by incorporating a predefined set of inclusion criteria.

1. The studies must report a clear distinction between use and non-use of ACEi/ARB as the two-comparator arms. If the studies provided data separately for ACEi or ARB then we intended to analyse the impact on outcomes independently. However, in the absence of non-segregation of data between ACEi or ARB, analysis will be performed for the combined use of ACEi and ARB.
2. Studies must include patients with SARS-CoV-2 infections only.
3. Severity of infection, death and hospitalisation was the primary outcomes of interest.
4. In the process of data analysis any additional outcome of importance could be included after arriving at a consensus.
5. All patients in the active-treatment arm must have been treated with ACEi/ARB.

We excluded all data presented in the form of commentaries, review articles, or case reports. Any data unrelated to SARS-CoV-2 infection were also excluded.

### 2.2 Process of study selection

The study was conceptualised by JJM. The article screening process was performed in a cyclical manner to make the process more robust. In phase one SG and BS conducted the web search jointly with mutual consultations. Any dispute was resolved on the basis of a mutually acceptable consensus. The inclusion and exclusion criteria were devised by JJM & KKG. Once the whole study selection process was completed, the team performed the same search in reverse order, with JJM & KKG doing the web-based screening and BS & SG formulating the selection criteria.

### 2.3 Study quality assessment

Quality of individual studies were assessed using the Cochrane collaboration tool using random sequence generation, allocation concealment, blinding of participants and personnel, blinding of outcomes assessment, incomplete outcome data, selective reporting and other bias as assessment attributes. Publication bias was assessed using funnel plots.

### 2.4 Statistical analysis

Analysis was conducted on a pooled population of 1842 patients identified with confirmed coronavirus infection from the six citations, using the comprehensive meta-analysis (CMA) software version 3, Biostat Inc., Englewood, NJ, USA. Heterogeneity was assessed using the Cochrane Q and Higgin’s I^2^ test and publication bias was assessed by funnel plots. [1–9] Depending on the presence or degree of heterogeneity (<45% low, 45–75% moderate and > 75% high) and the study characteristics, a fixed or random effect model to assess the effect size was selected. Where relative risk or odds ratio were not reported odds ratio was calculated from the reported events using Medcalc statistical software, © 2020 MedCalc Software Ltd, Ostend, Belgium.

## 3.0 Results

This meta-analysis includes patients from six studies with confirmed SARS-CoV-2 infection, and compares the outcomes among 423 patients receiving ACEi/ARB with 1419 patients who did not receive ACEi/ARB. Bean et al analysed patients who were on ACEi only, and Liu et al, on ARB only. The other studies did not segregate between patients on either ACEi or ARB. Hence, we have pooled the data and conducted this meta-analysis on the combined effect of ACEi/ARB.

The odds for risk of severity of disease was analysed from data extracted from all the six citations. The data from the study by Bean et al was included only for the analysis of the effect of ACEi/ARB on the risk of severity of disease as the odds ratio for this outcome in patients on ACEi was clearly mentioned in their publication; we could not include their data on effect on death since this was not reported separately as an outcome. (20). Similarly, data from the study of Liu et al (12) did not report on death separately. As such, the odds for death was analysed from the data extracted from the studies of Yang et al, Zeng et al, Zhang et al, and Li et al (17,19, 22). The odds for risk for hospitalisation was analysed from the data extracted from the studies of Yang et al (19) and Zeng et al (17) only, since the other four studies did not report on this outcome.

In view of the numerous terminologies used to report various end points in the six studies include in this analysis, we chose to assess the effect of ACEi/ARB on the risk of severity of disease. The data on death in the four studies was homogeneous as was the data on risk for hospitalisation among the two studies. The baseline characteristics of the studies are compiled in table 1. [Table 1]

**Tabel 1.**

| Study | Drug (n) | Non-ACEi/ARB (n) | Age in years | Odds ratio of Death | Odds ratio of severity of disease | Odds ratio of Hospitalisation | Additional information |
| --- | --- | --- | --- | --- | --- | --- | --- |
| Bean et al<sup>20</sup> | ACEi (37) | 168 | 63 ± 20 | NA | OR 0.42 (CI 0.14-1.00), p=0.058. | NA | NA |
| Liu et al<sup>18</sup> | ACEi/ARB (12) | 34 | 65.2 (Mean) | NA | OR=0.21, 95% CI: 0.05-0.85, p=0.03. | NA | NA |
| Yang et al<sup>19</sup> | ACEi/ARB (43) | 83 | 66 (Median) | OR 0.32 (CI 0.067 to 1.511, p=0.15) | OR 0.35 (CI 0.109 to 1.090, p=0.0699). | OR 0.95 (CI: 0.433 to 2.075, p=0.89) | Significant reduction in:<br>1. CRP (11.5 [4.0-58.2] vs 33.9 [5.1-119.2]; p=0.049.<br>2. Procalcitonin: 0.061 [0.044-0.131] vs 0.121 [0.052-0.295]; p=0.008 |
| Zeng et al<sup>17</sup> | ACEi/ARB (28) | 47 | 60 (Mean) | OR 0.65 (CI 0.116 to 3.577, p=0.62). | OR 2.46 (CI 0.939 to 6.449, p=0.07) | OR 0.57 (CI 0.178 to 1.812, p=0.34). | No difference in RT-PCR positivity |
| Zhang P et al<sup>21</sup> | ACEi/ARB (188) | 940 | 64 (Median) | OR 0.36 (CI 0.16-0.80, p=0.01) | OR 0.38 (CI 0.16-0.89, p=0.02) | NA | NA |
| Li J et al<sup>22</sup> | ACEi/ARB (115) | 247 | 66 (Median) | OR 0.57 (CI: 0.37-0.88, p=0.01) | OR: 0.62, 95% CI: 0.31-1.23, p=0.17) | NA | NA |

### 3.1 Effect of use of ACEi/ARB in patients with COVID-19 on risk of severity of disease (Figure 2)

There was a 38% statistically non-significant reduction in the odds of risk of severity of disease in patients with COVID-19 in the ACEi/ARB group compared to those not on ACEi/ARB (95% CI: 0.31–1.23, I^2^=70.36). There was moderate degree of heterogeneity.

**Figure 2.**
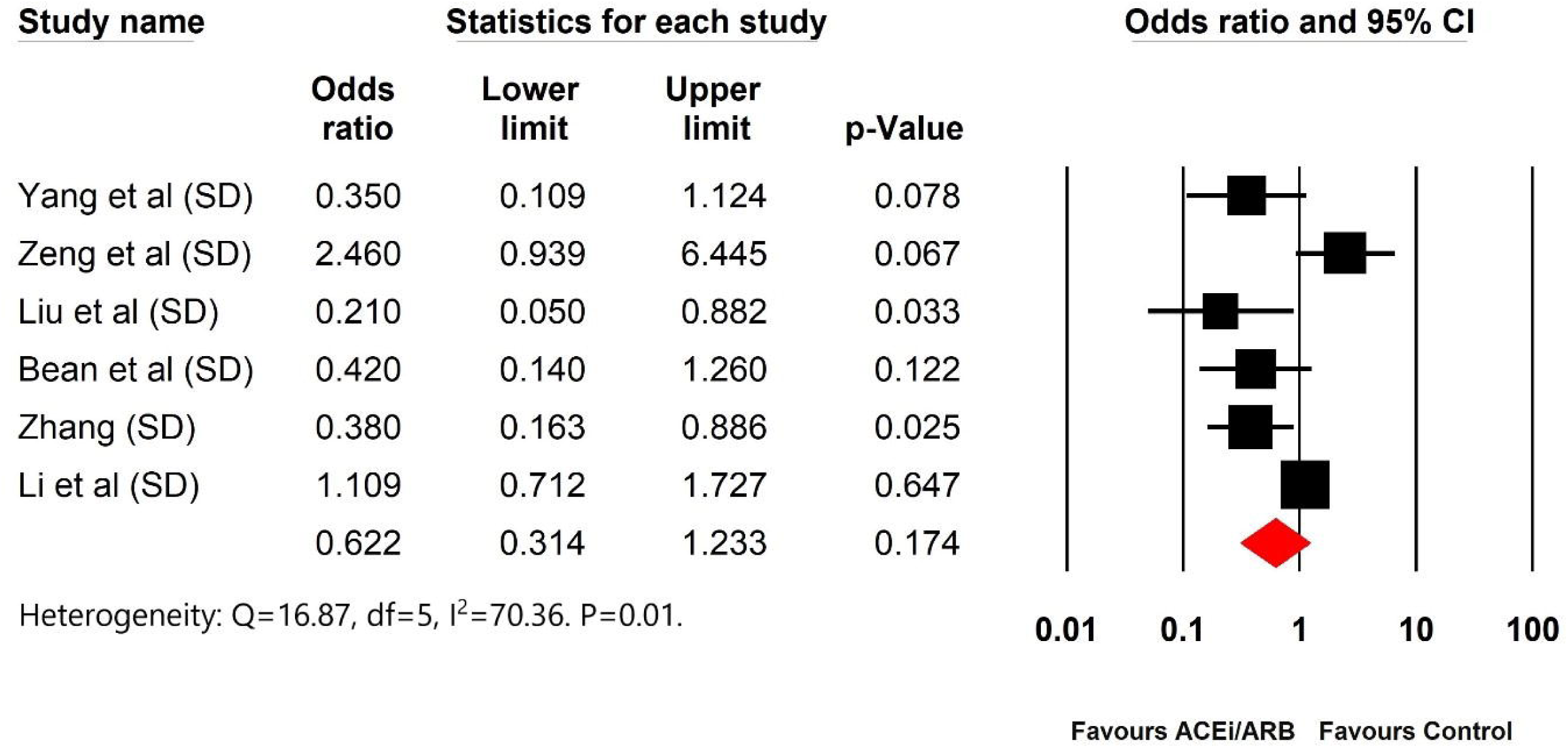

### 3.2 Effect of use of ACEi/ARB in patients with COVID-19 on death (Figure 3)

There was a statistically significant 43% (95% CI: 0.37–0.88, I^2^=0.000) reduction in the odds of death in the patients on ACEi/ARB when compared to those not on ACEi/ARB. There was negligible heterogeneity.

**Figure 3.**
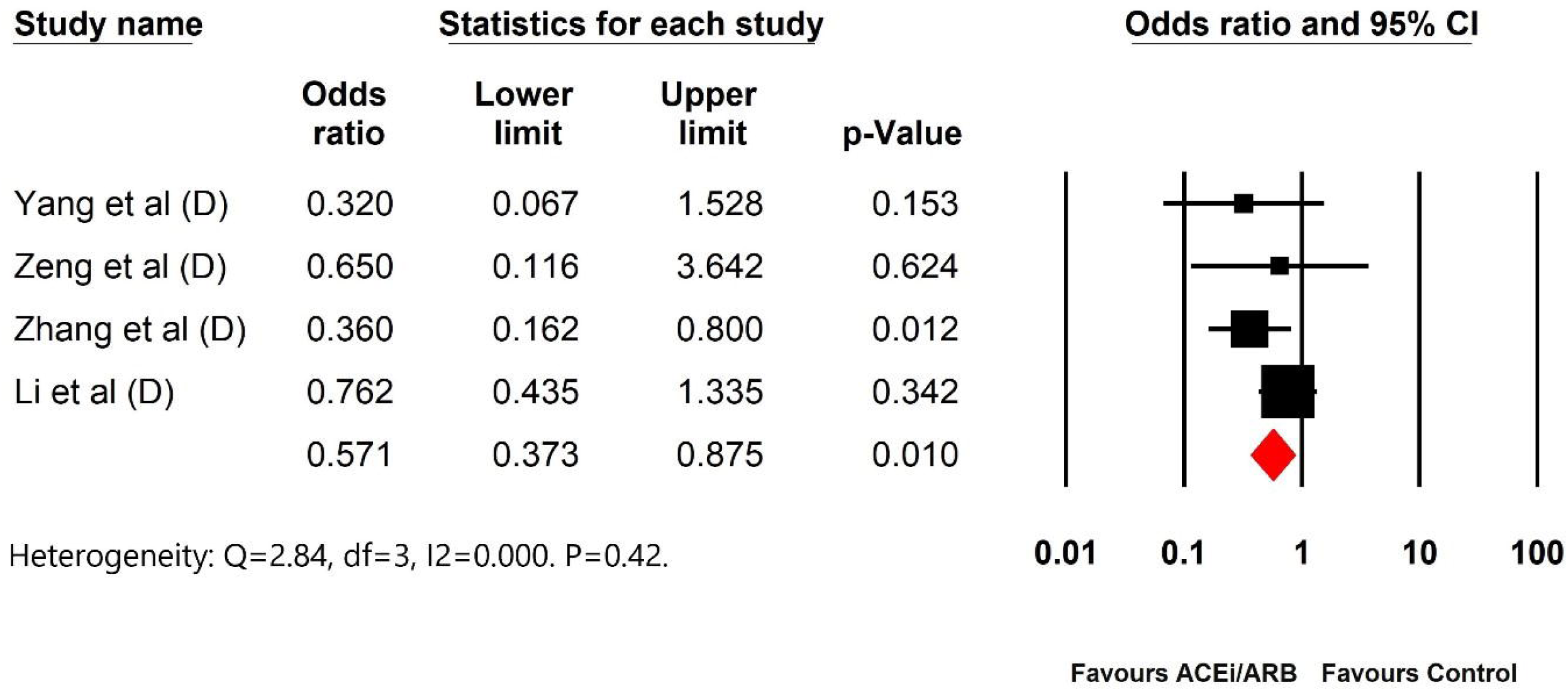

### 3.3 Effect of use of ACEi/ARB in patients with COVID-19 on risk of hospitalization

There was a statistically non-significant 19% (95% CI: 0.42–1.55, I^2^: 0.000) reduction in the odds of risk of hospitalisation in the patients on ACEi/ARB compared to those not on ACEi/ARB. There was negligible heterogeneity. (Figure 5)

**Figure 5.**
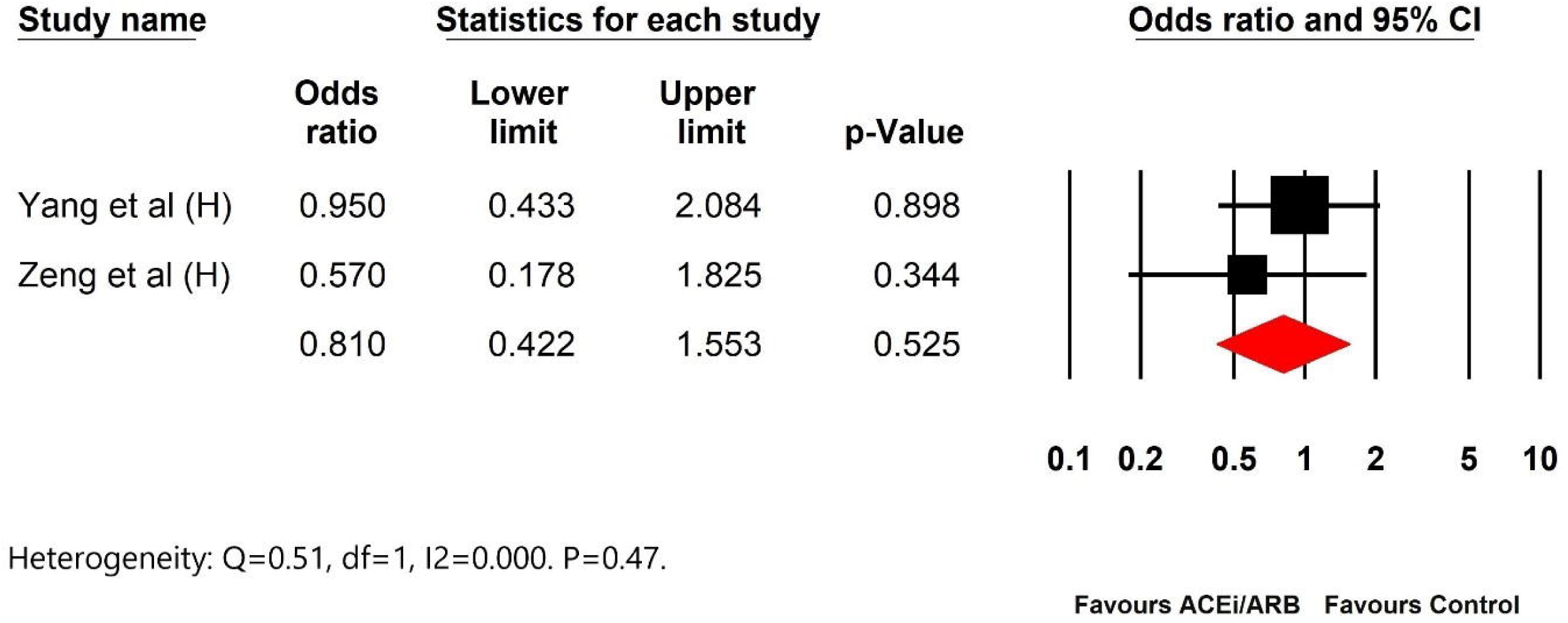

## 4.0 Discussion

Speculation over the deleterious effects of ACEi/ARB in patients with COVID-19 stems from the fact that SARS-CoV-2 uses the ACE2 receptor to enter target human cells (5). A few preclinical studies have suggested that RAAS inhibitors increase ACE2 expression, raising concerns regarding their safety in patients with COVID-19 (9). This thought was seemingly further strengthened by a retrospective study on hospitalised patients with COVID-19 in Wuhan, China, which reported that hypertensive patients on ACEi/ARB had an increased tendency to develop severe pneumonia with SARS COV 2 infection compared to those not on ACEi/ARB (54% vs. 32% respectively; p = 0.055) (17). This created confusion amongst patients and their health care workers since ACEi/ARB are an essential component of treatment not only in many individuals with hypertension but also in those with diabetes, coronary artery disease, heart failure and chronic kidney disease; sudden withdrawal of these medications in these patients might cause harm (22). However, findings from the same study showed that hypertensive patients with COVID-19 who were on ACEi/ARB had lesser hospitalisation (18% vs. 28%), lesser number of deaths (7% vs. 11%), and a higher number of discharges (75% vs. 61%) when compared to those not on ACEi/ARB; however, these observations failed to reach statistical significance due to the small number of patients analysed (17). In another study from China on elderly hypertensive patients with COVID-19, treatment with ARB was associated with a significant reduction in the risk of disease severity (OR=0·343, 95% CI 0·128–0·916, p=0·025) (18). A study by Yang et al. reported that compared to patients on non-ACEi/ARB antihypertensives, those on ACEi/ARB had a significant reduction in inflammatory markers, namely C-reactive protein (11.5 [ACEi/ARB] [4.0–58.2] v s. 33.9 [non ACEi/ARB] [5.1–119.2]; p=0.049) and procalcitonin (0.061 [ACEi/ARB] [0.044–0.131] vs. 0.121 [non ACEi/ARB] [0.052–0.295]; p=0.008), (19) consistent with previously described anti-inflammatory properties of ACEi/ARB (23, 24). This study also revealed a much lower proportion of critical patients (9.3% vs. 22.9%; p=0.061), and a lower death rate (4.7% vs. 13.3%; p=0.216) in those receiving ACEi/ARB compared to those not on ACEi/ARB; however, these differences failed to reach statistical significance (19).

In a study from the United Kingdom, Bean et al. reported that in patients with COVID-19, those treated with ACEi had a lower rate of death or transfer to a critical care unit [OR 0.29 CI 0.10–0.75 p <0.01], after adjusting for age, gender, and comorbidities (hypertension, diabetes mellitus, ischaemic heart disease and heart failure) compared to those who were not on ACEi/ARB (20). Recently, Zhang et al. reported a large multicentre retrospective study on 1128 hospitalised hypertensive patients with COVID-19; there was a significant reduction in all-cause mortality (adjusted HR, 0.30; 95%CI, 0.12–0.70; P = 0.01) in patients treated with ACEi/ARB compared to those treated with other anti-hypertensive medications, even after adjusting for a large number of variables (21). This study also demonstrated that use of ACEi/ARB was associated with lower risk of septic shock when compared to those not on ACEi/ARB (adjusted HR, 0.36; 95% CI, 0.16–0.84; P = 0.01) (21). In a recently published retrospective analysis of hypertensive patients from China, Li J et al. have reported that there was no difference in severity of disease (32.9% vs 30.7%; P = 0.645) or mortality (27.3%vs 33.0%; P = 0.34) among the hypertensive patients on ACEIs/ARBs when compared to those who were not on ACEi/ARB. (22)

The six studies described above, which have individually reported on the effect of ACEi/ARB in patients with COVID-19, are limited by the small number of patients and varied outcomes reported that makes it difficult to ascertain the effect of ACEi/ARB in patients with COVID-19. Ours is the first meta-analysis reporting the effect of ACEi/ARB on death and severity of disease in patients with COVID-19. We noted that patients with COVID-19 who were on ACEi/ARB had a 38% non-significant reduction in the odds for risk of developing severe disease [OR 0.62 (95% CI: 0.31–1.23), I^2^=70.36] when compared to those not on ACEi/ARB. The odds for risk of hospitalisation was also reduced by 19% (95% CI: 0.42–1.55, I^2^: 0.000) among patients with COVID-19 on ACEi/ARB compared to those not on ACEi/ARB, which failed to reach statistical significance. Most importantly, our meta-analysis of the four studies that have reported on death, reveals a statistically significant 43% reduction [OR 0.57 (95% CI: 0.37–0.88, I^2^=0.000) p=0.01] in the odds of death among patients with COVID-19 on ACEi/ARB compared to those not on ACEi/ARB, with negligible heterogeneity of data. The statistically significant benefits seen in our meta-analysis in the odds of death with use of ACEi/ARB in patients with COVID-19, together with a trend in reduction of severity of disease and risk for hospitalization, strengthens the recommendations from European Societies of Cardiology (25), Heart Failure Society of America, American College of Cardiology, American Heart Association (26) and International Society of Hypertension (27) to continue ACEi/ARB in patients with COVID-19. The reduction in the odds of death and the trend in the risk of developing severe disease and hospitalization is not only reassuring concerning the safety of ACEi/ARB in patients with COVID-19 but also point towards the urgent need for an adequately powered RCT to confirm these benefits. Till such time, our meta-analysis adds to current knowledge confirming not only the safety of ACEi/ARB in patents with COVID-19 but also possible benefit in terms of reduction in death.

These findings are in keeping with the results of a meta-analysis of three studies with 70346 patients with non-COVID pneumonia, where use of ARB was associated with a significant reduction in the risk of pneumonia-related morbidity (OR=0·55, 95% CI; 0·43–0·70, p<0·01) and mortality (OR=0·55, 95% CI; 0·44–0·69, p<0·01) (18). In another retrospective study on 313 elderly patients, it was noted that the relative risk of developing a new pneumonia of mixed aetiology was higher in patients who were not on an ACEi when compared to those who were receiving ACE inhibitors [18% vs. 7%; RR 2·65 (95% CI 1·31– 5·35, p=0·007)] (28). lt would appear that ACEi/ARB have a protective effect on pneumonia related pulmonary injury, which may be of benefit in patients with COVID-19 since the SARSCoV-2 has a predilection for the lower respiratory tract.

Our meta-analysis has a few limitations. The six studies included for analysis used different primary and secondary end-points that led to a moderate degree of heterogeneity in the reported results; we circumvented this by restricting our meta-analysis to clinically relevant end-points of death, hospitalisation and severity of disease, which resulted in negligible heterogeneity in our analysis on hospitalisation and death. The number of patients in the individual studies were small and there were various confounders, which were partly addressed by pooling of the data from the individual studies. Last, but not the least, due to a very small number of patients in the studies that reported outcomes independently in patients with COVID-19 on an ACEi or ARB, we were unable to assess the independent effect of ACEi or ARB on death and severity of disease in patients with COVID-19.

The strength of our meta-analysis lies in the fact that in an evolving infectious disease pandemic with acute consequences, where generation of evidence at the earliest is crucial to guide management, the best way to increase the predictability of the question in focus is to perform pooled analysis, till results of a randomized controlled trial (RCT) becomes available. By pooling data from multiple small observational studies with multiple inherent confounding factors, and by performing a meta-analysis, we have largely circumvented the problems associated with the individual studies. This meta-analysis, highlighting the safety and possibly beneficial effects of ACEi/ARB, shall help physicians use ACEi/ARB in patients with COVID-19 till further information becomes available. Secondly, since, the parameters assessed and the outcomes analysed varied enormously between the six studies, we analysed clinically relevant end-points including death, risk of hospitalisation, and severity of illness.

## 5.0 Conclusion

In conclusion, to our knowledge, this is the first meta-analysis on the effect of ACEi/ARB in patients with COVID-19 on clinically relevant end-points. We report that patients on ACEi/ARB, who develop COVID-19, do not have an adverse outcome; more importantly, within the limitations of a meta-analysis of retrospective observational studies, there was not only a trend in reduction of hospitalization and severity of disease, but also a statistically significant reduction in death. The findings of this meta-analysis reinforce the recommendations issued by the various agencies to continue ACEi/ARB in patients with comorbidities who contract COVID-19.

## Data Availability

All data used available online.

## References

1. Wu Z, McGoogan JM. Characteristics of and important lessons from the coronavirus disease 2019 (COVID-19) outbreak in China: summary of a report of 72314 cases from the Chinese Center for Disease Control and Prevention. JAMA. 2020;323(13):1239–1242

2. Wu C, Chen X, Cai Y, et al. Risk factors associated with acute respiratory distress syndrome and death in patients with coronavirus disease 2019 pneumonia in Wuhan, China. JAMA Intern Med. Published online March 13, 2020. doi:10.1001/jamainternmed.2020.0994

3. Zhou F, Yu T, Du R, et al. Clinical course and risk factors for mortality for mortality of adult inpatients with COVID-19 in Wuhan, China: a retrospective cohort study. Lancet 2020; 395(10299):1054–1062.

4. Severe Outcomes Among Patients with Coronavirus Disease 2019 (COVID-19) — United States, February 12–March 16, 2020. MMWR Morb Mortal Wkly Rep 2020; 69: 343–346

5. Hoffmann M, Kleine-Weber H, Schroeder S, et al. SARS-CoV-2 cell entry depends on ACE2 and TMPRSS2 and is blocked by a clinically proven protease inhibitor. Cell 2020; 181: 271–280.

6. Tipnis SR, Hooper NM, Hyde R, et al. A human homolog of angiotensin-converting enzyme. Cloning and functional expression as a captopril-insensitive carboxypeptidase. J Biol Chem. 2000; 274: 33238–33243.

7. Vickers C, Hales P, Kaushik V, et al. Hydrolysis of biological peptides by human angiotensin-converting enzyme-related carboxypeptidase. J Biol Chem. 2002; 277: 14838–14843.

8. Hamming I, Timens W, Bulthuis MLC, et al. Tissue distribution of ACE2 protein, the functional receptor of SARS coronavirus. A first step in understanding SARS pathogenesis. J Pathol 2004; 203: 631–637

9. Ferrario CM, Jessup J, Chappell MC et al. Effect of angiotensin-converting enzyme inhibitor and angiotensin II receptor blockers on cardiac angiotensin-converting enzyme 2. Circulation 2005; 111: 2605–2610.

10. Campbell DJ, Zeitz CJ, Esler MD et al. Evidence against a major role for angiotensin converting enzyme-related carboxypeptidase (ACE 2) in angiotensin peptide metabolism in human coronary circulation. J Hypertens 2004; 22: 1971–1976.

11. Luque M, Martin P, Martell N, et al. Effects of captopril relate to increased levels of prostacyclin and angiotensin (1–7) in essential hypertension. J Hypertens 1996; 14: 799–805.

12. Yang P, Gu H, Zhao Z, et al. Angiotensin-converting enzyme 2 (ACE2) mediates influenza H7N9 virus-induced acute lung injury. Sci Rep 2014; 4: 7027.

13. Liu Y, Yang Y, Zhang C, et al. Clinical and biochemical indexes from 2019-nCoV infected patients linked to viral loads and lung injury. Sci China Life Sci 2020; 63: 364–374.

14. Gu H, Xie Z, Li T, et al. Angiotensin-converting enzyme 2 inhibits lung injury induced by respiratory syncytial virus. Sci Rep 2016; 6: 19840.

15. Kuba K, Imayi Y, Rao S, et al. A crucial role of angiotensin converting enzyme 2 (ACE2) in SARS-coronavirus-induced lung injury. Nat Med 2005; 11: 875–879.

16. Kassiri Z, Zhong J, Guo D, et al. Loss of angiotensin-converting enzyme 2 accelerated maladaptive left ventricular remodelling in response to myocardial infarction. Circ Heart Fail 2009; 2: 446–455.

17. Zeng Z, Sha T, Zhang Y et al. Hypertension in patients hospitalized with COVID-19 in Wuhan, China: A single-center retrospective observational study. medRxiv preprint doi: https://doi.org/10.1101/2020.04.06.20054825

18. Liu Y, Huang F, Xu J, et al. Anti-hypertensive Angiotensin II receptor blockers associated to mitigation of disease severity in elderly COVID-19 patients. medRxiv 2020.03.20.20039586; doi: https://doi.org/10.1101/2020.03.20.20039586.

19. Yang G, Tan Z, Lou Z et al. Angiotensin II Receptor Blockers and Angiotensin-Converting Enzyme Inhibitors Usage is Associated with Improved Inflammatory Status and Clinical Outcomes in COVID-Patients With Hypertension.. medRxiv preprint doi: https://doi.org/10.1101/2020.03.31.20038935

20. Bean D, Kraljevic Z, Searle T, et al. Treatment with ACE-inhibitors is associated with less severe disease with SARS-COVID-19 infection in a multi-sile UK acute Hospital Trust. medRxiv 2020.04.07.20056788; doi: https://doi.org/10.1101/2020.04.07.20056788

21. Zhang P, Zhu L, Cai J et al. Association of Inpatient Use of Angiotensin Converting Enzyme Inhibitors and Angiotensin II Receptor Blockers with Mortality Among Patients With Hypertension Hospitalized With COVID-19. Circ Res. DOI: 10.1161/CIRCRESAHA.120.317134

22. Li J, Wang X, Chen J et al. Association of renin-angiotensin system inhibitors with severity or risk of death in patients with hypertension hospitalized for coronavirus disease 2019 (COVID-19) infection in Wuhan, China. JAMA Cardiol. (2020) doi:10.1001/jamacardio.2020.1624

23. Montecucco F, Pende A, Mach F. The Renin-Angiotensin System Modulates Inflammatory Processes in Atherosclerosis: Evidence from Basic Research and Clinical Studies. Mediators of Inflammation 2009, doi:10.1155/2009/752406

24. Dandona P, Dhindsa S, Ghanim H. et al. Angiotensin II and inflammation: the effect of angiotensin-converting enzyme inhibition and angiotensin II receptor blockade. J Hum Hypertens 21, 20–27 (2007). https://doi.org/10.1038/sj.jhh.1002101

25. European Societies of Cardiology. Position statement of the ESC Council on Hypertension on ACE-inhibitors and angiotensin receptor blockers. March 13, 2020. Available at: https://www.escardio.org/Councils/Council-on-Hypertension-(CHT)/News/position-statement-of-the-esc-council-onhypertension-on-ace-inhibitors-and-ang.

26. HFSA/ACC/AHA statement addresses concerns re: using RAAS antagonists in COVID-19. March 17, 2020. Available at: https://www.acc.org/latest-in-cardiology/articles/2020/03/17/08/59/hfsa-acc-aha-statement-addresses-concerns-re-using-raas-antagonists-in-covid-19.

27. A statement from the International Society of Hypertension on COVID-19. March 16,2020. Available at: https://ish-world.com/news/a/A-statement-from-the-International-Society-of-Hypertension-on-COVID-19/

28. Sekizawa K, Matsui T, Nakagawa T. ACE inhibitors and Pneumonia. Lancet 1998, 352: 1069

